# A Novel Hybrid Classical- Quantum Network to Detect Epileptic Seizures

**DOI:** 10.1101/2022.05.18.22275295

**Authors:** Mustafa Sameer, Bharat Gupta

## Abstract

**Background:** Machine learning (ML) has paved the way for scientists to develop effective computer-aided diagnostic (CAD) systems. In recent years, epileptic seizure detection using Electroencephalogram (EEG) data and deep learning models has gained much attention. However, in deep learning networks, the bottleneck is a large number of learnable parameters.

**Method:** In this study, a novel approach comprising a 1D-Convolutional Neural Network (CNN) model for feature extraction followed by classical-quantum hybrid layers for classification purpose has been proposed. The proposed technique has only 745 learning parameters, which is the least reported to date.

**Result:** The proposed method has achieved a maximum accuracy, sensitivity, and specificity of 100% for binary classification on the Bonn EEG dataset. In addition, the noise robustness of the proposed model has also been checked. To the best of the author’s knowledge, this is the first study to employ quantum machine learning (QML) to detect epileptic seizures.

**Conclusion:** Thus, the developed hybrid system will help neurologists to detect seizures in online mode.

## 1. Introduction

Epilepsy is a chronic neurological disorder, and around 1% population of the world is affected by it. Unprovoked recurrent seizures are the cause of epilepsy. Seizures are characterized by a sudden rush in the behavior of electrical pulses originating in the brain [1]. Clinicians are using Electroencephalogram (EEG) recordings to diagnose epilepsy. The visual examination of EEG is tedious, time-consuming, and error-prone. These factors led various researchers to develop an effective automated seizure detection system.

A vast number of studies are reported using hand-engineered features followed by machine learning techniques. Boonyakitanont et al. [2] reported an extensive survey of different feature extraction techniques used for epileptic seizure detection. Time-frequency analysis has been widely used on EEG time-series data for extracting the features [3]. Statistical features and different entropy variants are some of the major features used by various authors [4], [5]. The major disadvantage in hand-engineered techniques is it requires a specialist to select optimum features.

Deep learning eliminates the problem of selecting the best features and gives better results on huge amount of data. Deep neural networks (DNNs) usage has been increased tremendously in disease diagnosis, particularly neurological disorders [6]. The ability to extract features from the data in a sequential manner makes these networks more beneficial than conventional machine learning techniques. Shoeibi et al. [7] has presented a detailed survey on various deep learning approaches applied to epileptic seizure detection. Acharya et al. [8] proposed a 13 layer 1D-CNN model on the Bonn dataset. They have achieved 88.67% accuracy for three classes in 150 epochs. In another study, a pyramidal 1D-CNN approach has been presented to distinguish between seizures and non-seizures to reduce learning parameters [9]. Mahfuz et al.[10] has transformed EEG data into time-frequency images using Short-time Fourier transform and Continuous wavelet transform and then applied to different CNN models. Xu et al.[11] have presented a hybrid model consisting of both CNN and LSTM layers.

The papers mentioned above give good detection accuracy but on the adjustment/sacrifice with the complexity of the model, which makes them unaffordable or unnecessary for practical usages. The complexity of a deep learning model can be attributed to the number of filters, number of layers, smaller strides, and their combinations. The transformation of one-dimensional EEG data into two dimensions incorporates an additional stage and loss of important information. Generally, deep learning models use GPU to perform complex operations and reduce the time taken for computation.

Recently QML has taken an edge over its classical counterpart as it utilizes the computational ability of quantum computers. Shor algorithm is a perfect example of the above statement, which outlines the most efficient algorithm for the same task [12]. The fundamental idea of quantum computers is to break through the barriers that limit the speed of existing computers by harnessing the physics of subatomic quantum particles. Quantum computer has an advantage over classical computers in performing complex tasks faster as former utilizes the property of superposition and entanglement [13], [14]. Quantum bits, or qubits, are the basic units of information in quantum computing [15]. Qubit is represented as, |*ψ* ⟩ = *α* | 0⟩ + *β* |1⟩ with (*α*; *β* ∈ ℂ and | 0⟩, |1⟩ in the two-dimensional Hilbert space *H*^*2*^) [16]. Quantum computers are composed of quantum logic gates, which acts on qubits to change their state. All quantum circuits are implemented as a sequence of unitary operations using quantum gates [17]. Hence, QML algorithms can be designed as a quantum circuit to solve any specific problem.

### 1.1 Motivation and Contribution

Healthcare data which consists of electronic health records, information from clinical trials, disease registries, is growing at a compound rate of 36 percent yearly. Simultaneously, the number of healthcare consumers is increasing in using technology in their day-to-day lives. It has been proven that quantum technology is exponentially faster than classical computers [18]. Google reported a quantum processor that can perform a specific task in 200 secs that would take the world’s best supercomputer 10,000 years to complete [19]. The same results can be produced using simpler quantum models from the same data. Henderson et al. [20] show the advantage/ potential benefits of Quantum convolution neural networks over purely classical CNN model on MNIST dataset. The data in a quantum computer are represented in the superposition of quantum states, so it is easier to analyze the hidden patterns than classical computers [21]. However, current quantum computers have less than 100 qubits. Existing quantum computers have the greatest challenge of decoherence which causes the collapse of superposition states that contain vital information of qubit manipulations [22]. Therefore, to design machine learning algorithms that can fit into present-day quantum computers is a challenging task.

In this paper, the authors have designed a combination of classical and quantum circuits to classify the EEG signals for epileptic seizure detection. The advantage of hybrid approach is it overcomes the existing difficulties of quantum computers and allows the model to perform better. The major contributions of the present work are as follows:

1. A novel 1D-CNN model having only 703 parameters (least complexity to date) has been proposed to extract the features from EEG signals.
2. First time a hybrid classical-quantum classifier was implemented to detect epileptic seizures.
3. Noise robustness analysis was performed using hybrid classifier.

The paper is structured as follows: In Sect. 2, work done by researchers to detect diseases using quantum computing is presented. In Sect. 3, the Bonn EEG dataset, feature extraction, and classification block are described. In Sect. 4, hybrid classifier operation is explained, followed by simulation results obtained using Pennylane [23] framework. In Sect. 5, a discussion is presented between the proposed methodology and previous studies. Finally, in Sect. 6, we conclude this paper. Figure 1 shows the different stages of the proposed methodology, and each block is detailed in the next section.

**Fig. 1.**
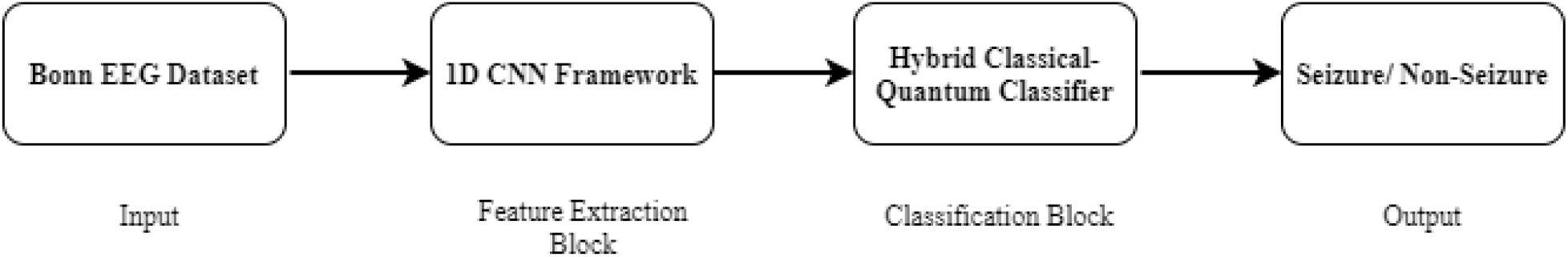
Different stages of the proposed methodology

## 2. Literature review on disease diagnosis using Quantum Computing

Quantum machine learning can revolutionize the diagnosis, imaging, treatment, and population health [24]. In the past two years, some researchers have applied quantum techniques to diagnose different diseases. A quantum framework has been presented for breast cancer detection in [25]. The classical-quantum transfer learning method has been implemented on IBM quantum computer for the detection of COVID-19 [26]. A 2-qubit hybrid quantum model has been presented to detect skin cancer in [27]. A quantum deep learning framework has been presented to detect diseases using phoniatrics biomarkers in [28]. In [29], authors have discussed a quantum perspective for osteoarthritis classification. In this paper, the authors have proposed a novel quantum approach to the regime of a hybrid classical-quantum algorithm to detect epileptic seizures.

## 3. Methodology

### 3.1 Dataset

The hybrid approach was implemented using the EEG signals collected from the Bonn University, Germany open-source database [30]. According to the standardized electrode placement scheme, the EEG signal recordings were recorded, i.e., 10–20 system, at a sampling rate of 173.61 Hz. The Bonn database consists of five different groups (denoted as Z, O, N, F, and S), and each group contains 100 segments. Each segment is of 23.6 seconds. Groups Z were collected from five mentally fit persons keeping eyes open. Group O was recorded during eyes closed from same subjects. Groups F and N were measured during the seizure-free interval (interictal state) acquired from the epileptogenic zone and hippocampal formation of the opposite hemisphere of the brain, respectively. Group S segments were recorded during the ictal state (seizure activity) of subjects.

### 3.2 Feature Extraction

#### Convolutional Neural Network (CNN)

The CNN network automatically acquires knowledge of EEG signals from the data as compared to a hand-engineered approach where features are selected by a specialist. CNN comprises convolutional layer (Conv), pooling layers, batch normalization layer, and fully connected layers. Conv layer has filters that detect different patterns of EEG signals. For feature extraction, small number of filters (kernels) are used. The number of filters is increasing in order, i.e., a small number of kernels or filters at low layers and a high number of filters at higher layers.

Pooling layers are used to reduce the volume of feature maps, which reduces the number of parameters of the model. Two types of pooling layers are used in the feature extraction model (a) Maximum Pooling and (b) Global Average Pooling.

Typically, in the last of the sequential CNN model, a fully connected layer connects every neuron in one layer to every neuron in another layer. It accepts flattened output from the convolutional layers. Dropout is used in CNN to prevent overfitting. In recent years, the number of layers is getting deeper, which has given rise to a large number of parameters. The recent emergence of making CNN deeper has given rise to a very large number of parameters that add to its complexity. To take care of a number of parameters (complexity), we have used a novel classical-quantum approach which drastically reduces the parameters. Table 1 shows the architecture of the proposed 1D CNN model.

**Table. 1.**
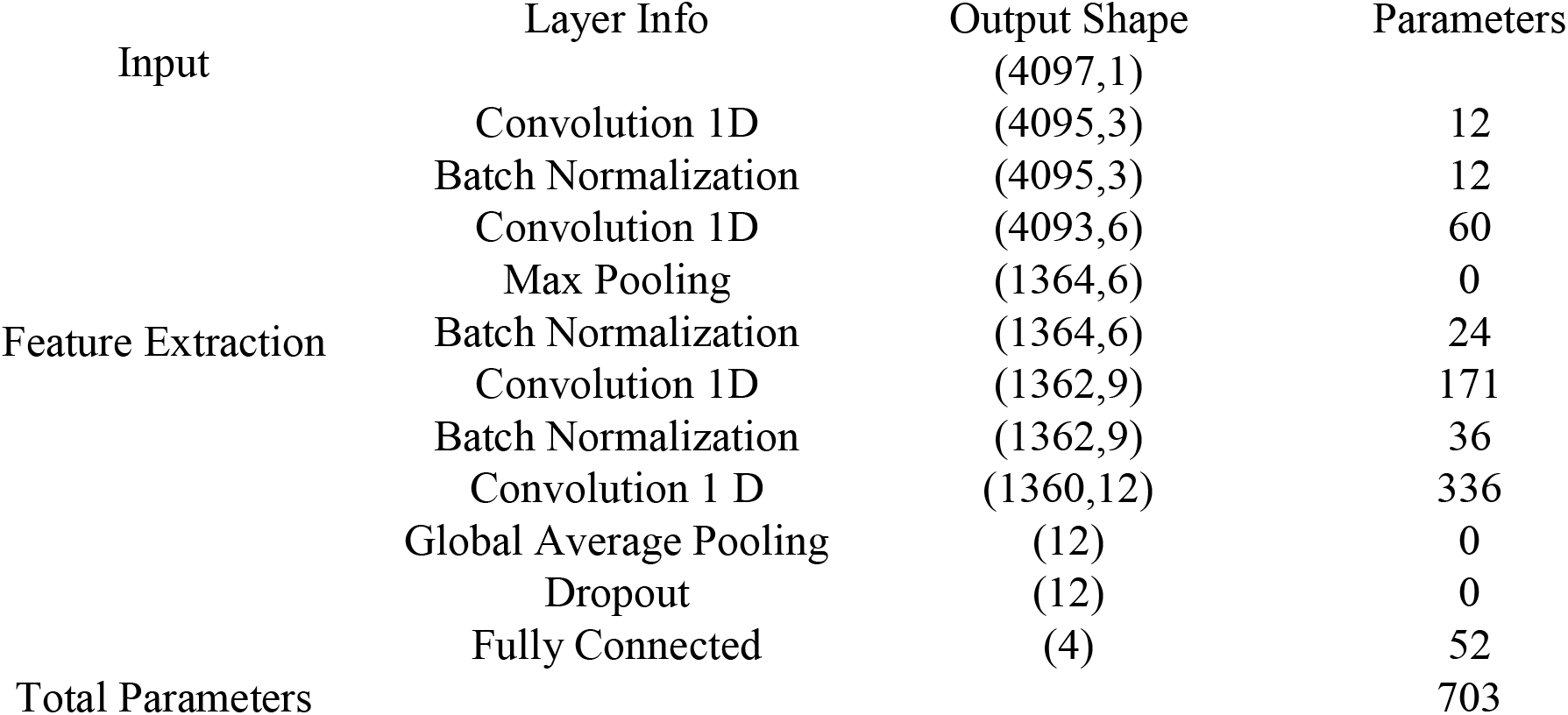
The specifications of the proposed 1D CNN model for feature extraction

### 3.3 Classification

A three-layered hybrid classifier has been used for detection purpose. Figure 2 shows the schematic of a hybrid classical-quantum classifier. The classifier consists of three layers which are as follows:

**Fig. 2.**
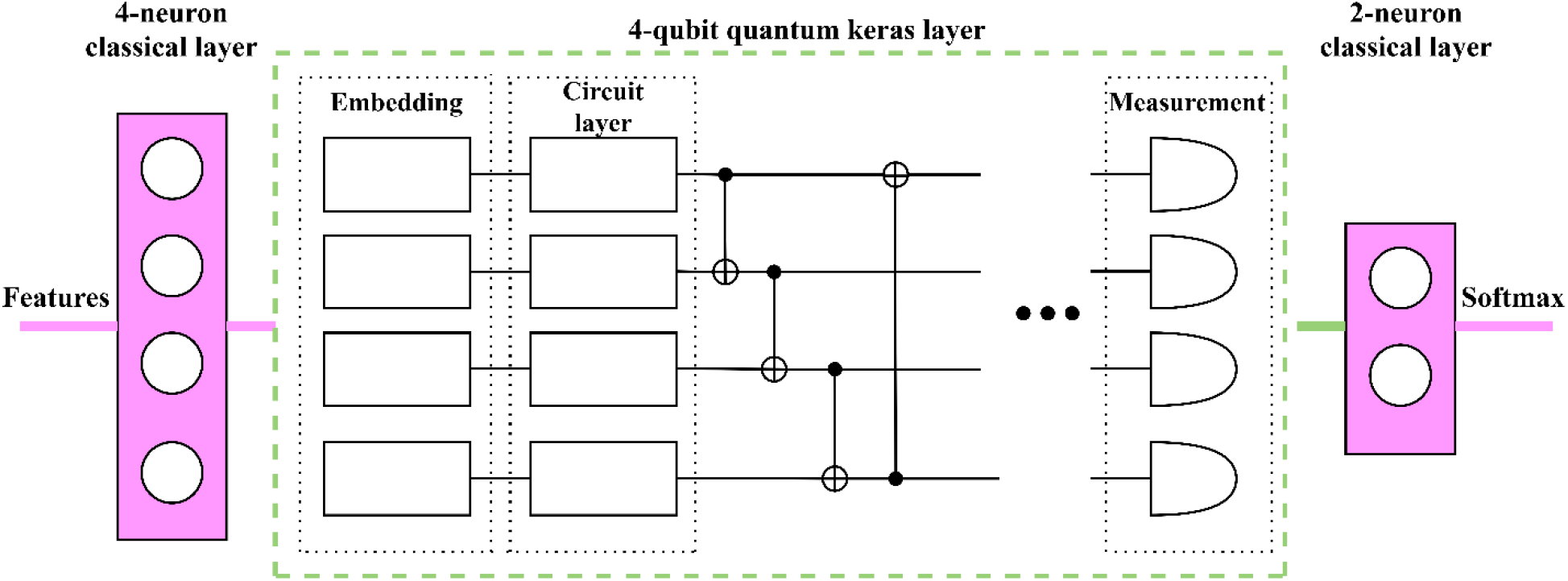
A general schematic of hybrid classical-quantum classifier

1. **4-neuron fully connected classical layer** – It takes the features extracted from the CNN as the input. It is just a regular layer of neurons same as in a neural network.
2. **Quantum layer:** It consists of different building blocks of quantum computing. Quantum computing effectively utilizes the potency of quantum mechanics such as superposition, entanglement, and interference for computing [31]. First part of the layer comprises an angle embedding layer that converts the classical bits into qubits. The second part of the layer is entanglement of the circuit, which is achieved with the help of CNOT gates to entangle the data [32]. The circuit layer performs quantum operations, i.e., rotation gates. It transforms the state of a qubit from one to another. Finally, from the measurement layer, we get the expected value of the qubit state. This 4-qubit quantum layer is implemented on the `default.qubit` simulator in Pennylane. The `qml.qnn.KerasLayer`, module simply wraps this quantum layer into a layer that’s compatible with TensorFlow and Keras, TensorFlow is able to classically optimize the network as if the layer was a classical one. The gradient for the quantum part of the network is calculated by different means depending on the device used, while all other gradients are calculated classically by TensorFlow. So, in short, training the weights is handled by TensorFlow as if it was all classically, while the quantum layer handles all the “quantum stuff” and returns classical outputs and gradients to the optimizer.
3. **N-neuron fully connected classical layer –** The output of the quantum layer is fed to this layer. N is determined by the number of classes. For binary classification, N is 2, and for three classes it is 3.

#### Softmax function

The output of the fully connected layer is fed to the softmax activation function. It is used to turn probabilities into logic numbers. The architecture of the classifier is presented in Table 2.

**Table. 2.**
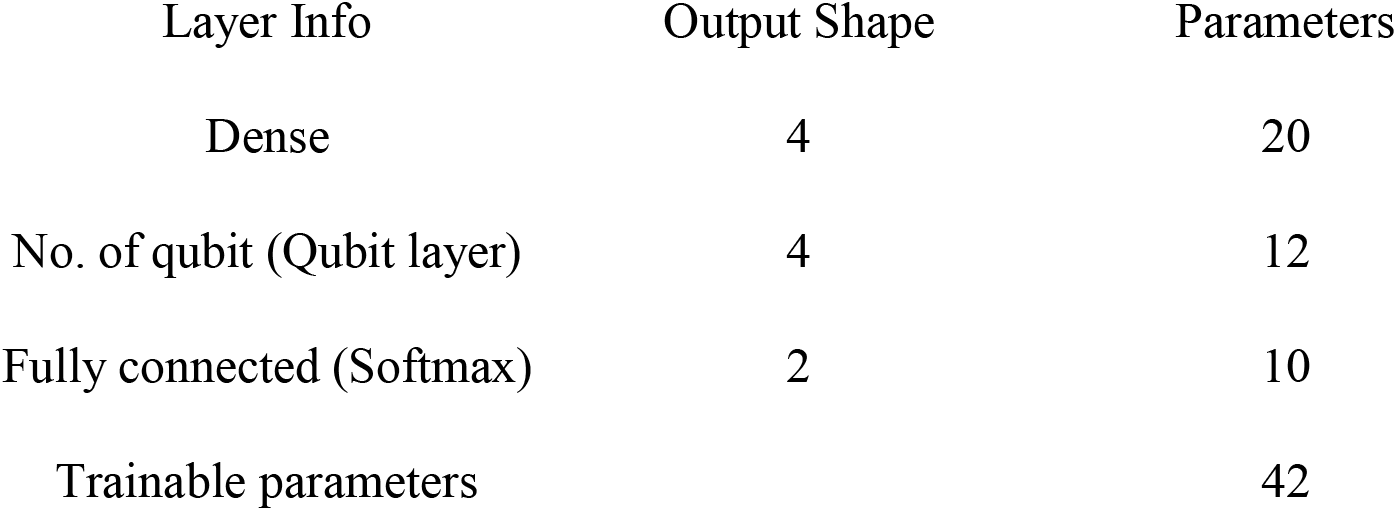
Description of the hybrid classifier

| Layer Info | Output Shape | Parameters |
| --- | --- | --- |
| Dense | 4 | 20 |
| No. of qubit (Qubit layer) | 4 | 12 |
| Fully connected (Softmax) | 2 | 10 |
| Trainable parameters |  | 42 |

## 4. Experiments and Results

In this section, the operation of the hybrid classical-quantum classifier has been explained, followed by the simulation results obtained on Bonn EEG dataset.

### 4.1 Details of hybrid classifier operation

The classifier consists of a mixture of units having classical and quantum computations. The entire hybrid classifier is implemented using Pennylane. In Pennylane, the quantum components are represented using an object called a Quantum Node or QNode. A quantum node consists of a quantum function and a device on which it executes. A quantum device or a quantum simulator is initialized using `qml.device` method for quantum operations in Pennylane with the number of qubits given as input to wires.

In this work, the quantum node has one layer and is programmed in three modules: Angle Embedding, Strongly Entangling circuits, and Measurement. Angle embedding encodes the classical information into rotations. The entanglement portion is achieved with the help of CNOT gates. The measurement operation is executed on each wire using `qml.exp.PauliZ(k)`, where k = 1, 2,…n is the label of wire. The quantum node is implemented on the `default.qubit` simulator. The QNode object is then transformed to a Keras layer by using `qml.qnn.KerasLayer(qnode, weight_shapes, output_dim= n_qubits)`. It creates a TensorFlow compatible tape, and then the model is trained with the Keras Adam optimizer. The QNode comes with a specific `diff_method=best` which is used to evaluate the gradient in the background for the given device. Softmax function is used to obtain the predicted labels. So what’s happening here is basically:

1. The weights are updated (classically) and supplied to the QNode function.
2. The QNode uses these exact values as parameters for applying whatever operations are used in the circuit.
3. The QNode returns an output (after applying the circuit operations) along with a calculated gradient.

### 4.2 Simulation Results

To evaluate the performance of the proposed model, the benchmark Bonn EEG dataset is used in this work. Eight binary classifications and one three-class classification have been performed. The data clusters (a combination of different groups) are divided into 70:30 ratios for training and testing purposes. The simulations have been performed using Tensorflow and Keras library. The four features are extracted from 1D-CNN model and fed to hybrid classical-quantum classifier. Early stopping monitor function has been utilized in the feature extraction from CNN. Batch size of 4 has been taken, and Adam is used as an optimizer with a learning rate of 0.001. Table 3 shows the classification performance of the proposed approach in terms of accuracy, sensitivity, and specificity. Data cluster S-Z obtained 100% results for all the three metrics only in 200 epochs. S-NF and S-ZONF have also achieved 100% sensitivity. S-O-F has achieved a sensitivity of 89.28%.

**Table. 3.** Classification measures of the proposed hybrid classical-quantum approach

| Data Cluster | Epochs | Accuracy | Sensitivity | Specificity |
| --- | --- | --- | --- | --- |
| S-Z | 200 | 100 | 100 | 100 |
| S-O | 200 | 98.33 | 96.67 | 100 |
| S-N | 500 | 96.67 | 100 | 93.33 |
| S-F | 300 | 91.67 | 96.67 | 86.67 |
| S-ZO | 1000 | 97.78 | 96.43 | 98.39 |
| S-NF | 1000 | 95.56 | 100 | 93.55 |
| S-ZONF | 800 | 92 | 100 | 90.24 |
| ZO-NF | 1500 | 89.17 | 85.29 | 94.23 |
| S-O-F | 1000 | 83.33 | 89.28 | 80.65 |

To check the robustness of the proposed model, it has been tested against the noise. For the noise analysis, polluted signals were generated by adding white Gaussian noise into the original signal. The noise status of the signal is monitored by the standard deviation (σ) of actual noise. In the experiments we chose three σ values i.e. 0.1, 0.3, 0.5. Figure 3 shows the radar plot for the accuracy and sensitivity obtained. It can be observed that in all data clusters, results are almost the same for different values of σ except S-O-F.

**Fig. 3.**
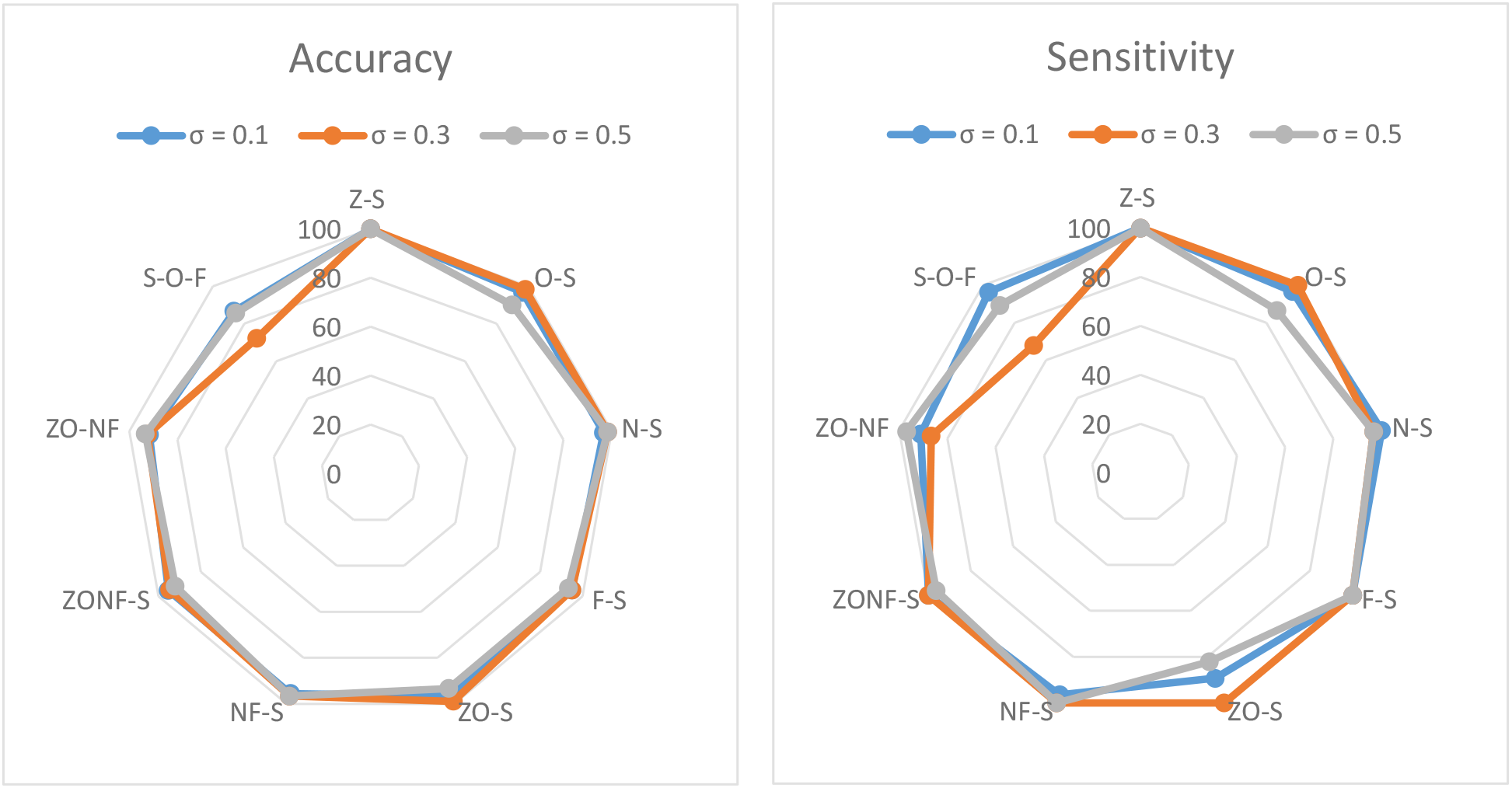
Noise analysis for different values of σ

## 5. Discussion

Many deep learning models have been reported for epileptic seizure detection. But their main disadvantage is high trainable parameters. As the parameters increase, the time complexity of the model also increases. Table 4 shows the comparison with other techniques between seizures and non-seizures. Thara et al. [33] reported a two-layer BiLSTM (Bidirectional Long Short Term Memory) model having 39549 parameters and achieved 99.08% accuracy. Segundo et al. [34] have achieved good accuracy, but the trainable parameters are 251297. Ansari et al. [35] have used the CNN model having 7600 parameters for the detection of neonatal seizures. The proposed hybrid technique has only 745 parameters, and the results are almost comparable. So it is one of the simplest models to date.

**Table. 4.** Comparison of proposed approach with other existing studies

| References | Methodology | Trainable Parameters | Accuracy (%) |
| --- | --- | --- | --- |
| [33] | Stacked BiLSTM | 39549 | 99.08 |
| [36] | Fourier + wavelet + Empirical Mode Decomposition + DNN | 251297 | 99.8 |
| [9] | Pyramidal 1D CNN | 8326 | 100 |
| [35] | CNN and Random Forest | 7600 | 77 |
| This study | CNN and Hybrid Classical Quantum classifier | 745 | 100% |

Quantum algorithms are still evolving. At this point, researchers can only do experiments. It is still not accessible for clinicians or health givers for monitoring purposes as quantum hardware is still developing. The main challenge here is determining the optimum number of layers and filters with the tradeoff between accuracy and complexity. Another area of optimization is to explore different encoding methods converting from bits to qubits.

## 6. Conclusion

A novel hybrid classical-quantum network has been presented in this paper to classify EEG epilepsy. A total of eight experiments has been performed on the benchmark Bonn EEG dataset. 1D-CNN of the least complexity has been utilized to extract the features. The extracted features have been fed to the hybrid classifier for classification. The sturdiness of the proposed technique has also been evaluated by adding Gaussian noise to the EEG signal. This is the first study to use quantum computing to detect epileptic seizures. QML is in its nascent stage, and authors have planned to optimize the present study.

## Data Availability

The dataset analyzed during the current study are available in the repository
https://repositori.upf.edu/handle/10230/42894

https://repositori.upf.edu/handle/10230/42894

## Data Availability

The dataset analyzed during the current study are available in the repository https://repositori.upf.edu/handle/10230/42894

## Funding

This work did not receive any grant from funding agencies in the public, commercial, or not-for-profit sectors

## Compliance with ethical standards

### Conflict of interest

The authors declare that they have no conflict of interest.

### Ethical approval

This article does not contain any studies with human participants or animals performed by any of the authors.

## Notes

### Competing Interest Statement

The authors have declared no competing interest.

### Funding Statement

This study did not receive any funding

### Author Declarations

https://repositori.upf.edu/handle/10230/42894

